# Federated penalized piecewise exponential model for horizontally distributed survival data: FedPPEM

**DOI:** 10.64898/2026.02.11.26346054

**Authors:** Nazmul Islam, Chongliang Luo, Jiayi Tong, Daniel A. Pollyea, Craig T. Jordan, Bradley Haverkos, Steven Bair, Andrew Kent, Grant Weller

**Affiliations:** RefinedScience, Aurora, Colorado; Division of Public Health Sciences, Department of Surgery, Washington University, St. Louis, Missouri; Department of Biostatistics, Bloomberg School of Public Health, Johns Hopkins University, Baltimore, Maryland; Department of Medicine, University of Colorado Anschutz Campus, Aurora, Colorado

**Keywords:** Federated penalized survival model, privacy-preserving algorithm, multi-site collaboration, horizontally distributed data structure, AML risk stratification

## Abstract

Cox proportional hazard regressions are frequently employed to develop prognostic models for time-to-event data, considering both patient-specific and disease-specific characteristics. In high-dimensional clinical modeling, these biological features can exhibit high collinearity due to inter-feature relationships, potentially causing instability and numerical issues during estimation without regularization. For rare diseases such as acute myeloid leukemia (AML), the sparsity and scarcity of data further complicate estimation. In such cases, data augmentation through multi-site collaboration can alleviate these problems. However, this often necessitates sharing individual patient data (IPD) across sites, which presents challenges due to regulatory barriers aimed at protecting patient privacy. To overcome these challenges, we propose a privacy-preserving algorithm that eliminates sharing IPD across sites and fits a federated penalized piecewise exponential model (FedPPEM) to estimate potential effects of clinical features using summary statistics. This algorithm yields results nearly identical to those from pooled IPD, including effect size and standard error estimates. We demonstrate the model’s performance in quantifying effects of clinical features and genetic risk classification on overall survival using real-world data from ∼1200 newly diagnosed AML patients across 33 U.S. sites. Although applied in AML context, this model is disease-agnostic and can be implemented in other diseases and clinical contexts.

## 1. Introduction

Survival models are extensively employed to assess the relationships among clinical features, biomarkers, and end-points, particularly accounting for censoring in clinical trials and drug development. The Cox proportional hazard (PH) model, in particular, has gained significant popularity in survival regressions due to its simplicity, interpretability, and the availability of software. However, in real-world high-dimensional settings, fitting a Cox-PH model for rare hematological disorders (e.g., acute myeloid leukemia (AML)) is challenging due to the sparsity of features and the limited number of patients. Moreover, other real-world data (RWD) challenges, such as the curse of dimensionality due to the abundance of molecular features, collinearity among features, missing data, patient heterogeneity, and treatment variations across sites, further complicate the task numerically. To address these issues, multisite collaboration that pools individual-level patient data (IPD) across sites is one potential solution. However, sharing IPD to facilitate such collaboration is often impractical due to institutional policies, patient privacy concerns, and/or regulatory barriers. Regulations such as the Health Insurance Portability and Accountability Act^1^ (HIPAA) in the United States or the General Data Protection Regulation^2^ (GDPR) in the E.U. prohibit the sharing of sensitive IPD across sites without strict protocols and data sharing agreements. Although de-identification or encryption techniques can enable IPD sharing to a certain extent, reverse engineering through cross-joining datasets may still expose personally identifiable information and thus threatening patient privacy. In recent years, privacy-preserving algorithms, such as federated learning (FL) techniques, have been extensively studied and explored across various domains. However, their application to clinical problems and real-world evidence (RWE) generation specifically in oncology remains sporadic and underdeveloped.

In the realm of federated statistical and machine-learning (S/ML) models, the use of survival data has gained a lot of attention recently. Different modeling techniques for vertical (i.e., data features) and horizontal (data subjects) survival data have been developed, and they are still in their early stages^3,4^ ^5^. A federated Cox-PH model can be horizontally fitted by calculating site-specific gradients exploiting the corresponding partial likelihoods and then averaging them across sites for a global representation. However, this straightforward aggregation does not accurately approximate the true global gradient because precise risk set calculations at each unique event time require data from all individuals, not just local ones. The One-shot Distributed Algorithm Cox model^3^ (ODAC) calculates the derivatives of the global likelihood function at each site; however, this approach necessitates the initial sharing of sensitive event times. To address these challenges, Westers et al^6^ proposed horizontal federated CoxPH models by transforming the survival modeling framework into a more manageable logistic regression through stacking. Andreux et al^7^ suggested discrete-time survival models by converting time-to-event data into a binary classification problem, treating event times as a covariate. However, while the stacking approach for estimation necessitates sharing patient-level event times across sites, this risk can be mitigated by employing the date shifting method. Nevertheless, the threat of patient re-identification remains significant for smaller sites, particularly when patients have extreme follow-up periods for a rare disease; furthermore, such random shifting may obscure temporal pattern of effects perturbing inference and validation. Additionally, computational challenges increase in high-dimensional real-world data settings, especially when biomarkers are sparsely distributed or correlated across sites. In a similar spirit, Miao et al^8^ fitted a vertically distributed penalized Cox model in a high-dimensional setting, which requires initially sharing the risk sets for all subjects with other sites to ensure event ordering. Such data sharing is not often feasible for horizontally distributed federated models as it requires enhanced coordination across sites. Furthermore, this may lead to unwarranted protected health information (PHI) exposure for both horizontally and vertically distributed models. An alternative federated method for fitting Cox-PH model on vertically distributed survival data coined as VERTICOX^9^ has been proposed to resolve data sharing issues. However, none of these approaches accommodate regularization and uncertainty quantification of coefficient estimates, which are crucial for inference in a high-dimensional setting.

In this article, we introduce a privacy-preserving FL algorithm tailored for horizontally partitioned survival data. structure^10^, Our proposed model approximates the non-separable CoxPH model by employing the piecewise exponential data (PED) structure, thus transforming it into a more manageable generalized linear modeling framework which is computationally more amenable. Specifically, this allows a separable loss function, enabling flexible modeling for high-dimensional data with both fixed and varying coefficients. The proposed model is accompanied with an L_2_-norm (ridge) penalty under the assumption of a Poisson distribution and is coined as the federated penalized piecewise exponential model (FedPPEM). The novelty of the proposed approach includes: 1) estimation using summary statistics without pooling any IPD (such as risk sets, event times, event ordering, event indicators, etc.) for retaining patient privacy, 2) introduction of regularization penalties to facilitate high-dimensional estimation, 3) uncertainty quantification and inference through summary statistics (local hessian) and bootstrapping, 4) communication efficiency, and 5) interpretability of effect sizes, flexibility in model specification, and computational effectiveness.

To illustrate the practical value of FedPPEM in a high-stakes, multi-institutional setting, we apply it to risk prediction and stratification in acute myeloid leukemia (AML). AML is a rare and aggressive blood disorder, with nearly 20,000 individuals diagnosed annually in the United States^11^. Detailed phenotypic and molecular data at diagnosis has revealed significant disease heterogeneity amongst AML patients. This, paired with the ever-expanding landscape of treatment options, has motivated prognostic modeling efforts to help pair each patient with the best possible treatment for them. Patient risk stratification and treatment-specific response prediction from the outset is crucial to achieve the best clinical outcomes. Furthermore, population-level stratification aids in determining patient allocation, selection, and enrichment for clinical trials during the pre-treatment phase, while also playing a significant role in predicting treatment response and making decisions about treatment changes, maintenance or other adjustments later in the disease course. As more sub-stratification of the disease occurs through more nuanced testing modalities, larger patient cohorts are required to gain statistically robust insights about these ever-smaller populations. To ensure sufficient sample size and events for analysis and generate robust evidence, multi-institutional collaborations are a necessity. Recently, several risk stratification models for AML have been newly developed to allow for more accurate prognostication in the context of treatment with venetoclax-based therapies, including the refined European Leukemia Network (rELN24)^12^ and refined risk model^13^ (RRM). In our study, we aim to quantify and validate these two AML risk models using the proposed FedPPEM method with RWD of 1192 de-novo AML patients treated with venetoclax and azacitidine (ven/aza) at 33 independent sites (University of Colorado (CU) and 32 external sites). In the Results section, a concise overview of the FedPPEM algorithm is presented, and its performance is assessed empirically using the multi-site data. We examine the combined data from these 33 institutions through standard penalized survival regression and compare the findings with those obtained from applying the proposed FedPPEM algorithm to the same dataset without sharing IPD. The FedPPEM method will pave the road for integrating more clinical sites into the research of AML and other rare diseases as well.

## 2. Results

We transformed the survival model into the PED structure where follow-up times are divided into a distinct grid of small intervals, within which the baseline hazard is assumed constant^14^. The selection of the number of knots for determining global time intervals was context-dependent; in our application, we opted for smaller time intervals during the early follow-up period as AML has a higher likelihood of relapse in early time-period, but a relatively lower relapse thereafter. The FedPPEM fits a generalized linear model (GLM) with a Poisson distribution assuming log link function in a high-dimensional setting adjusting for confounders. An L_2_-norm penalty was employed to ensure numerical stability, address collinearity among biomarkers and clinical attributes, and mitigate the risk of overfitting. The regularization term facilitates estimation by utilizing information from small sites within a high-dimensional framework, particularly when the number of covariates surpasses the number of subjects^15^. The coefficients characterize the association between biomarkers and events in terms of hazard ratio (HR). Unlike the pooled PEM via GLM, the federated model leverages the site-specific likelihood value, gradient vectors, and hessian matrices to iteratively optimize estimation over a grid of penalty values. Tuning parameters were empirically selected from a dense grid of pre-specified values that minimize the Akaike Information Criteria (AIC) or Bayesian Information Criteria (BIC). To reduce communication rounds between a site and the orchestrator, each site shared the necessary information (i.e., likelihood, gradient, and Hessian) associated with each penalty value with the orchestrator in a single round. The outer loop was executed to optimize parameter estimation using Newton-Raphson separately for a given penalty value, and this process was repeated over all pre-specified grid values of penalty term. Figure 1 illustrates the modeling steps, and further details of the algorithm are provided in the Methods section. Supplemental Figure 1 highlights the steps generating bootstrap-based confidence intervals via FL mechanism.

**Figure 1.**
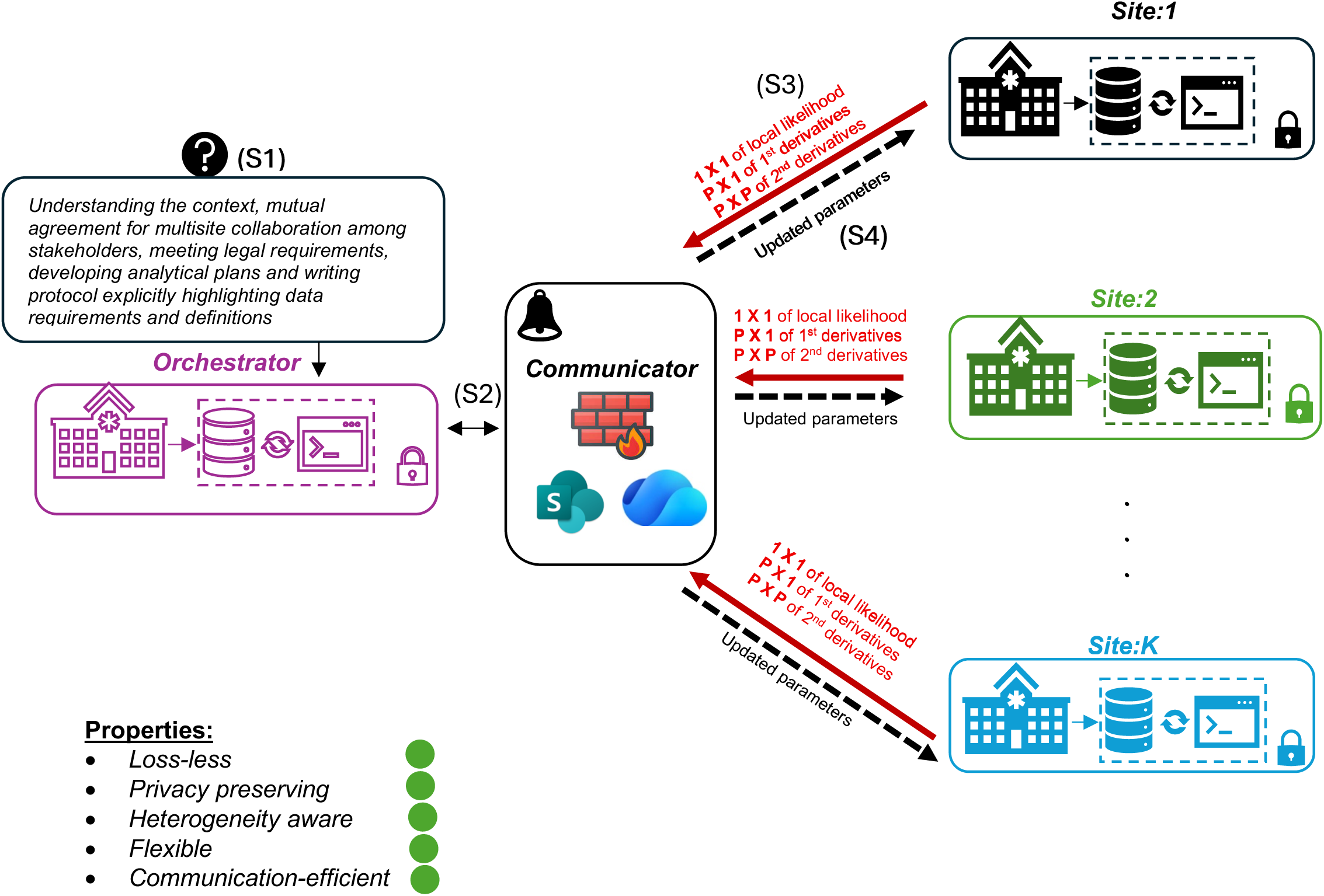
An overview of federated penalized piecewise exponential model framework. Steps are : (S1) standardizing, normalizing, and developing the protocol that to be shared with sites for data definitions, derivations, transfers and usage; (S2) sharing codes, initial values, knots, and grid of tuning parameter(s) with sites; (S3) enumerating site-specific summary statistics and sharing with the orchestrator; (S4) updating coefficient estimates iteratively and sharing with the sites to recompute summary statistics; (S5) running (S3-S4) iteratively till convergence or pre-specified number iteration is reached; (S6) sharing optimum parameters with participating sites.

We empirically demonstrated the statistical properties of the proposed method by quantifying the association of genetic feature-based risk stratification, which categorizes each patient as Adverse, Intermediate, or Favorable, while controlling for the effects of demographics (age, gender) and comorbidities (obesity, prior non-AML cancer, myelodysplastic syndromes (MDS), heart-disease, hypertension, hyperlipidemia, chronic obstructive pulmonary disease (COPD), hypothyroidism, coagulopathy, renal disease (CKD), venous thromboembolism (VTE)). For our empirical investigation, we utilized a horizontally distributed dataset comprising of AML patients treated with ven/aza across 33 unique sites. Figure 2 illustrates the distribution of patients (i.e., cases or mutations) showing varying feature distributions across these sites.

**Figure 2.**
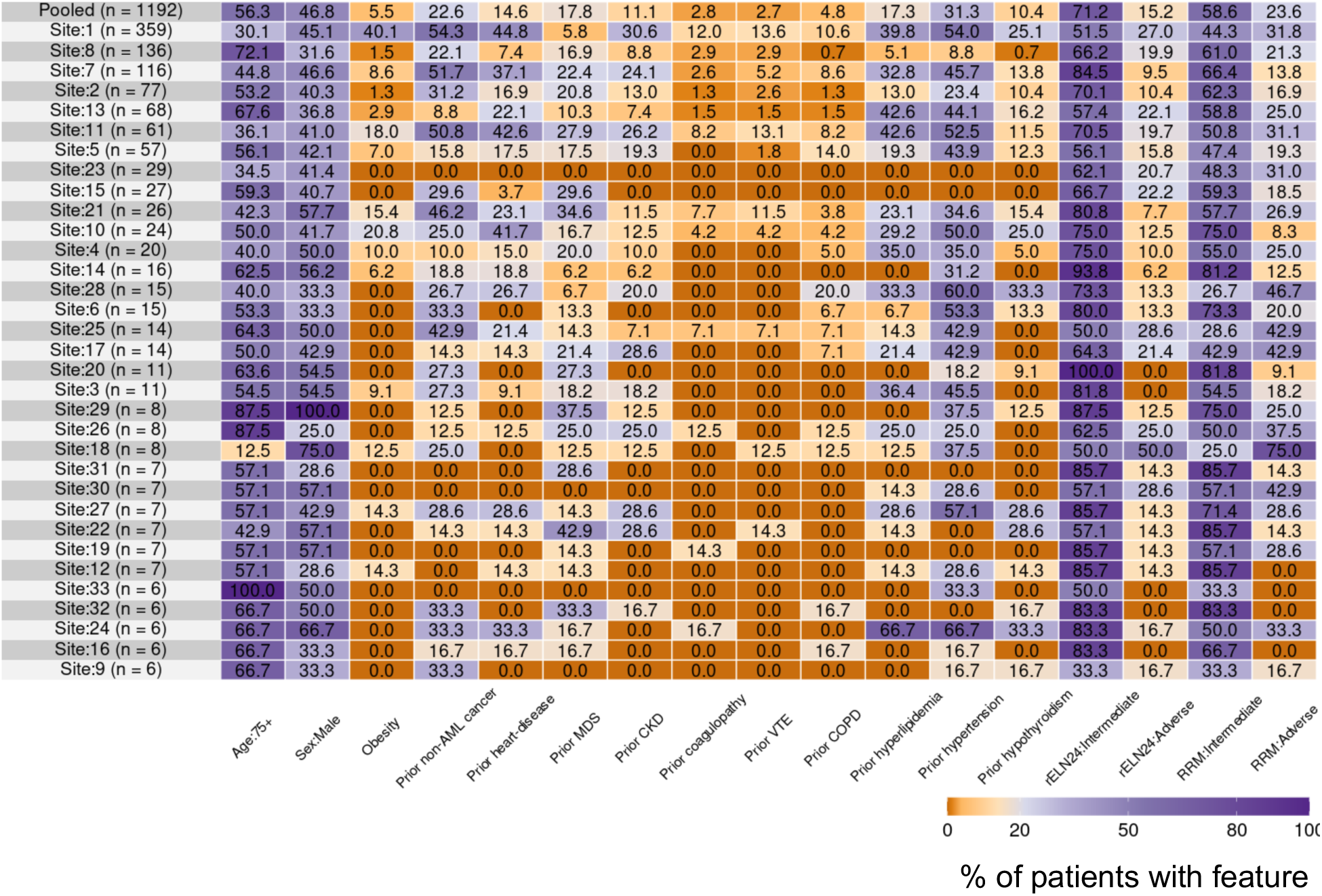
Distribution of covariates across 33 sites for 1192 patients. Reported in each cell are the proportion (%) of patients with each feature (i.e., mutated or “Yes”). Age is dichotomized as greater or equal to 75 years old. Color shed varying from purple to red highlights the variability across sites. Overall distribution of patients is highlighted at the top.

With access to all local datasets, we were able to compare numerical accuracy across different methods. For this comparison, we conducted a pooled analysis by merging all datasets, a federated analysis without sharing any IPD, and a random-effect meta-analysis by aggregating estimates of the local models. To examine the effects of the number of sites and the number of patients per site, we created three scenarios including: (a) sites with more than 5 patients and (b) sites with more than 20 patients, and (c) sites with more than 50 patients. The first condition permits greater diversity in patient mix, leading to a larger sample size and an increased number of participating sites (K = 33 vs 11 vs 7). Enabling small-site participation is important for producing RWE, as rural community hospitals often have a limited patient base, which typically precludes them from contributing to local knowledge generation and results in their frequent omission from clinical trials or studies.

In the top panels of Figure 3, the FedPPEM’s loss-less property is demonstrated, as the estimates match those from the pooled analysis for both the coefficient estimates and their corresponding standard errors in the scenario with more than 50 patients per site. The empirical results with more than 5 (Supplemental Figure 2) and 20 (Supplemental Figure 3) patients per site also yield similar outcomes. In contrast, the results of the random-effect meta-analysis reveal significant estimation discrepancies (Figure 3, bottom panels). This is expected, given that meta-analyses are highly sensitive to factors such as site size, patient heterogeneity, covariate distribution, and methodological differences. Notably, the meta-analysis with (a) and (b) above led to numerical issues and unstable parameter estimates.

**Figure 3.**
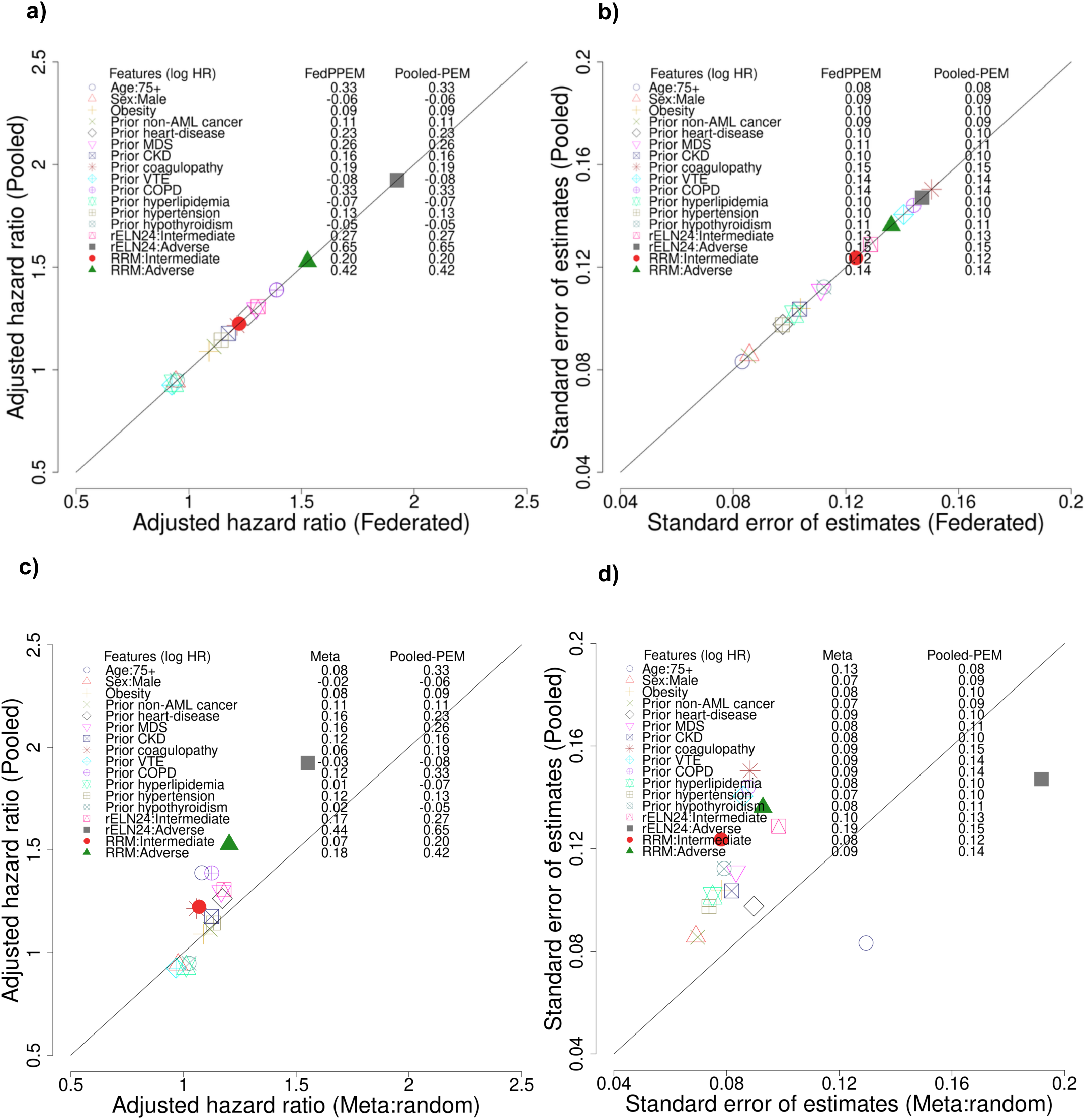
Comparison between federated and pooled estimates using FedPPEM and meta-analysis. Reported are (a) parameter estimates for federated versus pooled, (b) Corresponding standard errors, (c) estimates derived from random effect meta-analysis compared to pooled, and (d) associated standard errors. These results are based on data from 7 sites, each with more than 50 patients, totaling 874 patients in the pooled analytical set.

Both standard errors and bootstrapping were adopted for inference. To account for bias stemming from the regularization penalty of the FedPPEM, we used fractional random weight bootstrap (FRWB) approach to generate confidence intervals (CI) instead of using model-based standard errors using hessians; Supplemental Figure 4 compares the bootstrap percentile-based 95% CI’s lower and upper bounds derived from the pooled and federated models.

Figure 4 illustrates the adjusted hazard ratios (aHRs) for potential confounders with bootstrap-specific 95% CIs using 300 runs with respect to the conditions determined by the size of each participating site (a-c). Clinical characteristics such as age, previous CKD, heart disease, COPD, and hypertension are negatively associated with OS in all different scenarios, although the strength of these associations varies as the power and distribution of the patient characteristics change. Both risk categories, rELN24:Adverse (aHR 1.92; 95% CI 1.66-3.03) and RRM:Adverse (aHR 1.53; 95% CI 1.21-1.93), demonstrate negative associations with OS. However, the risk difference between the Adverse and Favorable groups is more pronounced for rELN24 compared to RRM. Here higher aHR values (Adverse vs Favorable and Intermediate vs Favorable) indicate greater differences in OS among the three groups suggesting better overall separation among survival curves of rELN24 compared to RRM. Unsurprisingly, the estimates from larger sites with more patients (N = 1192) yield better inferential outcomes with narrower confidence intervals. RRM estimates seem to have smaller variation relative to rELN24. From a clinical standpoint, these large validation studies of prognostic models help fine tune predictive models and increase the precision of patient-level treatment decisions.

**Figure 4.**
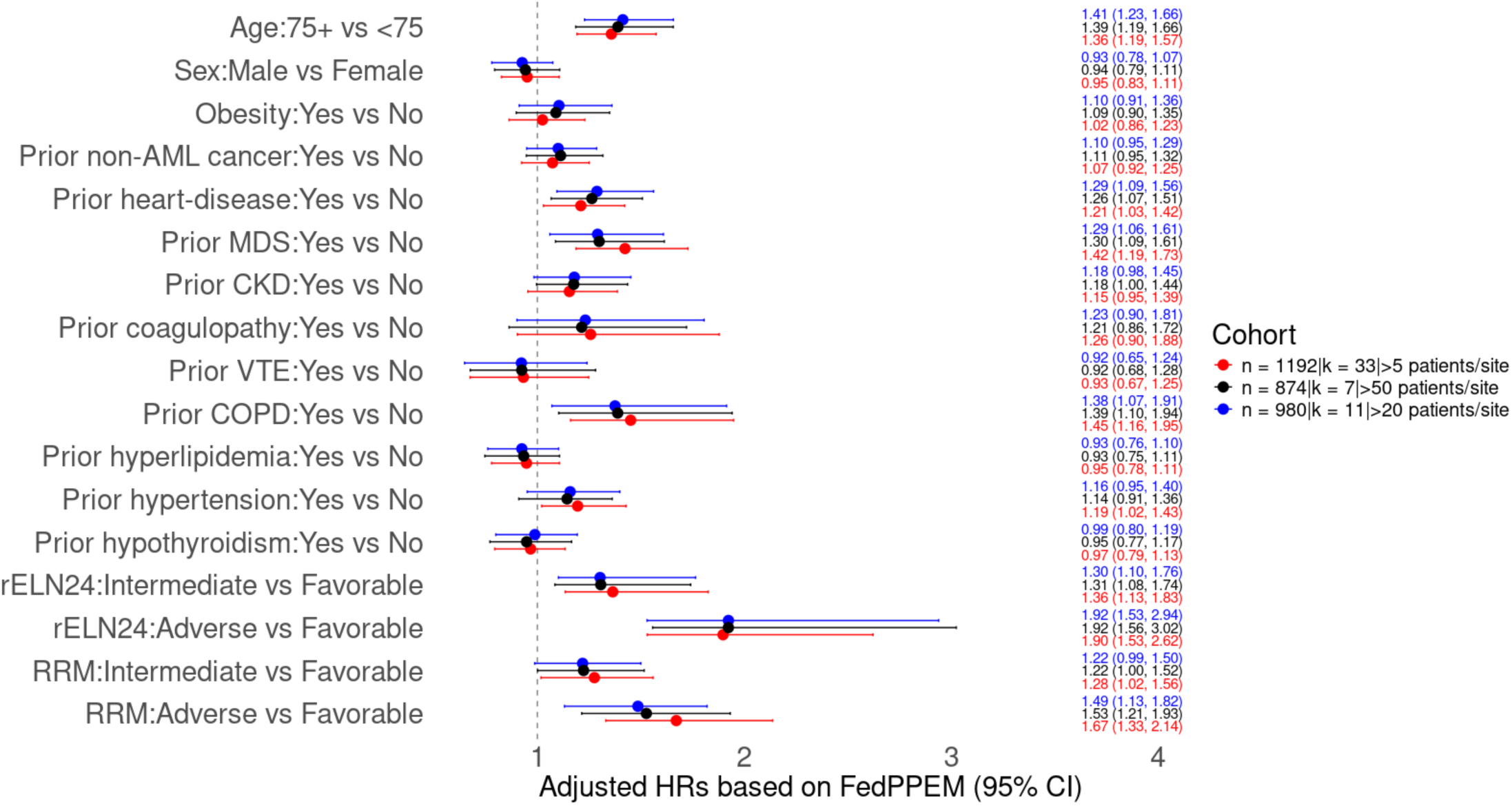
Forest plot of FedPPEM based adjusted hazard ratio. Reported are the estimates along with bootstrap based 95% confidence intervals for three analytical sets. (blue) N = 980 and K = 11 having more than 20 patients per site, (black) N = 874 and K = 7 having more than 50 patients per site, and (red) N = 1192 and K = 33 having more than 5 patients per site.

Figure 5 depicts the incremental improvement in numerical accuracy towards convergence with additional Newton-Raphson steps taken. Results are based on the analytical set including sites having more than 50 patients. Generally, FedPPEM requires approximately 12 communication rounds (steps) between a site and the orchestrator to reach convergence for an optimally selected ridge penalty value from a dense grid of 1e-5 to 1e5. However, the model attains results close to the optimal after just 8 communication rounds indicating the efficiency and robustness of the model; where all the differences in coefficients are within the range of (1%, 15%). Note that the convergence and computation time are influenced by the granularity of knots or time intervals; denser and smaller intervals necessitate more rounds of communication to achieve convergence. For ease of implementation, it is recommended to determine the number of knots based on the clinical context, pattern of experiencing events, and study objectives. We implemented the model using both 6 and 12 knots, finding that the coefficient estimates for confounding features were approximately similar, although the latter required more computational time.

**Figure 5.**
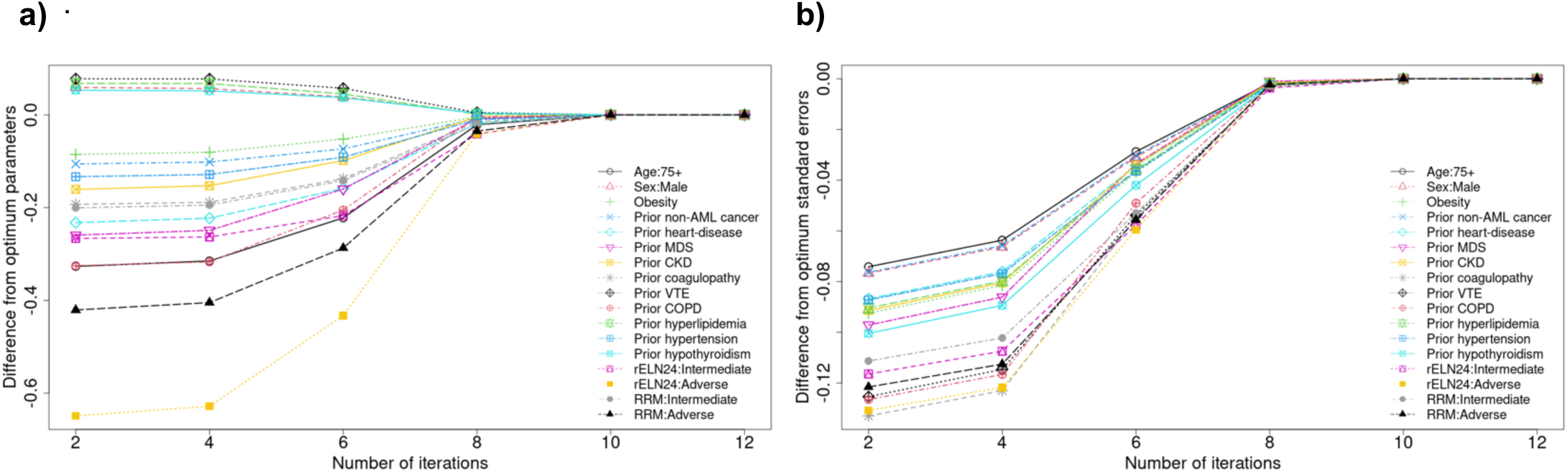
Convergence of FedPPEM estimates over different Newton-Raphson iteration steps. Results are based on the sites having more than 50 patients (N = 874 and K = 7) for (a) parameter estimates and (b) the corresponding standard errors.

## 3. Discussion

In this study, we introduced a federated penalized PEM tailored for horizontally distributed survival data, which demonstrates flexibility, maintains data integrity, and preserves privacy while optimizing model parameters through a communication-efficient method. The proposed model does not necessitate access to IPD, sensitive risk information, or event times which are securely stored at each respective site. Instead, it relies on site-specific summary statistics, including a scalar value, a gradient vector, and a second-derivative matrix, for estimation purposes at each round. The model exhibited superior numerical performance in both estimation and inference, highlighting its advantages in terms of stability and robustness compared to Meta-analysis. The PED structure simplifies the transition from time-to-event regression to Poisson regression, allowing for the use of flexible modeling approaches like GLM. The incorporation of regularization acts as a safeguard against overfitting and interdependencies among covariates in high-dimensional real-world data. Additionally, the process of generating confidence intervals via distributed FRWB ensures reliable inference for sparse datasets with limited sample sizes. Although the proposed FedPPEM was applied to AML datasets from CU and external collaborators, it can be adapted and utilized in other contexts (e.g., different survival endpoints or disease areas) without the loss of generalizability. Implementing such a disease-agnostic strategy can significantly enhance the ability to extract valuable clinical insights regarding the effects of biomarkers on survival endpoints, even for the rarest diseases. This approach addresses the immense challenges posed by small sample sizes scattered globally, all while ensuring the protection of patient privacy.

The findings validate the efficacy of both the rELN24 and RRM risk models, with the former showing a more pronounced separation between risk-based OS curves. This risk stratification can be considered a single-dimensional characteristic that encapsulates the impact of various genetic features (such as CYT, FISH, NGS, PCR, etc.), making it useful for initial patient selection, prognostic modeling, and treatment management decisions. Identifying factors linked to time-to-event, including age, heart disease, MDS, COPD, renal disease, and hypertension, highlights the necessity for comorbidity-specific risk modeling alongside genetic-based risk stratification.

There are several limitations and future directions to consider. A) The use of Poisson regression with PEM to approximate Cox-PH models assumes a constant hazard rate within each time segment, and the precision of this approximation is influenced by the granularity and size of the time intervals. Supplemental Figure 5 depicts subtle differences between the estimates of penalized Cox-PH model and penalized PEM. B) While the model is designed to enforce patient privacy, it has not been tested for comprehensive robust privacy-preserving techniques such as differential privacy or k-anonymity related to re-identification of patients via linking data to external data sources. C) In general, it is difficult to confirm whether there are overlapping patients across different sites; for example, the multicenter RWC includes patients from all locations across the U.S. In our application, we confirmed that there was no duplication of AML patients between the RWC and CU. However, such confirmation is not always possible when data comes from a large consortium or central database. D) Even though the protocol for normalization and standardization was set before the study began, consistently maintaining data definitions was not always feasible due to missing data and variations in test types across sites. Often, assumptions had to be made based on available data resulting in deviation from the protocol. For instance, the biomarkers used in generating RRM were defined slightly differently in the RWC compared to the CU due to missing features, different test types (NGS, PCR, FISH, CYT), and testing equipment; for more details, refer to Islam et al 2024. E) The site-specific files (i.e., matrix of Hessians) might be too large to share, and the corresponding matrix operation (e.g., inversion of a matrix) may present numerical challenges, especially for very high-dimensional single-cell omics datasets. Similarly, matrices for smaller sites with sparse local data may need transformation to convert them into positive definite matrices for numerical ease. F) The current method does not account for varying effects of features (i.e., non-linear effects), which may require incorporating polynomial or smoothing effects into the model and we defer this for future exploration. G) The proposed methodology is designed for association and inferential tasks and is not suitable for predicting individual risk, which may require hazard function estimation and non-linear machine/deep learning (ML/DL) based modeling strategies. H) The proposed model is not intended for estimating treatment effects within a causal modeling framework. However, the FedPPEM could be easily extended for causal ML models by integrating G-computation or inverse probability of weights with some modifications in the proposed steps. I) Note that the proposed model does not consider heterogeneity between sites. However, the model can be extended with some adjustments. One approach involves (i) incorporating a site-specific intercept and modifying the design matrices accordingly, (ii) introducing site-covariate interaction effects along with an additional ridge penalty term, or (iii) including a random effect for sites.

The proposed FedPPEM enables collaboration across multiple sites by allowing both small and large sites contributing to generate RWE. This approach promotes inclusivity by incorporating a diverse patient population from various sites spanning different countries and continents, leading to a more robust and dependable estimation of population-level parameters.

## 4. Method

### 4.1 Description of analytical datasets

The primary site (orchestrator) contains 359 adult patients with newly diagnosed AML patients treated with venetoclax/azacitidine between 2015 and 2024 in the University of Colorado (CU) cancer center. There are 32 secondary sites whose data is obtained from the Flatiron health. Even though we have access to these secondary sites’ datasets, for the development of FL model, the corresponding sites’ datasets were treated as locked meaning there is no sharing of IPD across sites. The Flatiron Health dataset is a licensed, de-identified dataset comprising patient information on individuals diagnosed with AML using ICD-9 and ICD-10 codes. These individuals had a minimum of two documented clinical visits between 2014 and 2023 and received venetoclax/azacitidine as a frontline therapy. During the study period, the de-identified data were sourced from approximately 280 cancer clinics across the United States, encompassing around 800 sites of care, primarily within community oncology settings. The data were de-identified and subject to obligations to prevent re-identification and protect confidentiality. For validation and illustration, sites having more than 5 patients were selected. All site-specific datasets were horizontally distributed containing patient-specific demographics, comorbidities, and AML diagnostic pathology features including next generation sequencing (NGS), cytogenetics (CYT), fluorescence in-situ hybridization (FISH), polymerase chain reaction (PCR), flow cytometric (FC), and composite mutations using various tests. Risk stratification models (rELN24 and RRM) were applied independently to these datasets to obtain patient-level risk classification utilizing the genetic variables observed at diagnosis. We imputed missing genetic features locally as imputation-by-mode. Alternatively, federated model-based imputation technique can also be adopted; the details of these methods are discussed elsewhere and beyond the scope of this paper.

### 4.2 Piecewise exponential model for survival data

Let *δ_i_* and 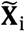 be the event indicator and *p*-dimensional covariate vector for the *i*-th subject respectively. Define *κ_i_* be the time-to-event for the i-th subject. Similarly, censoring time is denoted by *c_i_*. Denote the survival data ^1^ where *t_i_* = *min*(*κ_i_*, *c_i_*). The hazard function under the Cox-PH is written as 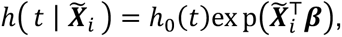 where ℎ_0_(. ) is unknown hazard function and ***β*** is a set of unknown parameters quantifying the association between features and survival end-points. We leverage the statistical equivalence between the Cox-PH model and the PEM assuming ℎ_0_(. ) is approximated as a constant over fine-grid of time intervals. We cast the model into a PEM framework utilizing the PED structure where the transformed design matrix is defined by ***X****_i_*. Define the PEM by

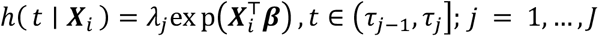

where *λ_j_* is constant within each discrete time intervals. Note that when intervals (*τ_j_*_-1_, *τ_j_*]’s are short and dense enough, this model approaches the Cox-PH model and yields nearly identical parameter estimates. The strength of the classical PEM is that multivariate survival analysis of time-to-event data can be performed using the algorithms designed to fit widely used GLM framework via maximizing the Poisson likelihood function assuming log link function^16,17^. Let the number of events for the *i*-th subject at the *j*-th interval be

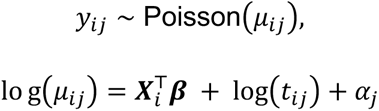

where *μ_ij_* is the mean number of events at the *j*-th interval, *log*(. ) is an offset term which is the interval length serves to account for time duration each subject is at risk in each interval, and *α_j_* = *log*(*λ_j_*) is log baseline hazard for the *j*-th interval.

### 4.3 Federated penalized piecewise exponential model

To fit penalized PEM, it is essential to expand and aggregate likelihood function by sites, a process made possible by the separability characteristic of the loss function based on the Poisson likelihood. In federated settings, each site retains its local data, sharing only summary statistics which are discussed in the next section along with the steps of fitting FedPPEM. Let *k* denote the number of sites where *k* = 1, …, *K*. Let ***n****_k_* be the number of subjects in each site, ***X****_k_* be the (***n****_k_* × *p*) design matrix of covariates with row being a (*p* × 1) vector of ***X****_ik_*′*S*, ***y****_k_* be the ***n****_k_* × 1 vector of *y_ijk_*′*S*. Define the offset term by ***o****_k_*. To adjust for collinearity among features, an L2-norm penalty was used resulting in a regularized Poisson GLM. The corresponding penalized likelihood function can be expressed as

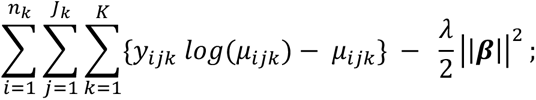

where 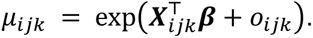

It has been shown that a federated algorithm can reconstruct the same global score and Hessian by exchanging summary-level information, which leads to estimators that asymptotically converge to the pooled penalized estimators through the Newton-Raphson scheme^18–21^, and in our algorithm described below, we have leveraged this concept.

### 4.4 Estimation of federated penalized piecewise exponential model

The estimation of parameters is described as below.

1. Each **site** performs standardization and normalization of features and impute any missing values according to the pre-defined protocol and shares maximum follow-up time. Any deviation to the protocol across sites is identified.
2. **Orchestrator** shares analytical codes (.R) for implementation, grid of potential tuning parameters ***λ***, and pre-specified global knots (e.g., 6 or 12) for time-interval which are selected based on clinical context.
3. Each **site** independently transforms survival response and design matrix into piecewise exponential data (PED) format and obtain ***X****_k_*, ***y****_k_*, and ***o****_k_* exploiting dataset with size *n_k_*.
4. Set initial parameters as ***β****_λ_*_,0_ = {0, …, 0}. For each value in ***λ***, each **site** shares the following unpenalized aggregated statistics with the **orchestrator**-

a. (*p* ×*1*) vectors of gradients 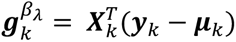 where ***μ****_k_* = exp(***η****_k_*) ; 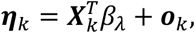
b. (*p* × *p*) matrices of hessians 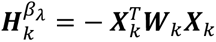 where ***W****_k_* = *diag*(***μ****_k_*),
c. Scalar values of likelihood 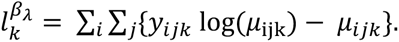
5. **Orchestrator** aggregates the site-specific shared data; 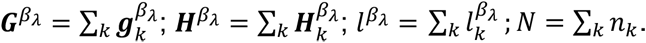
6. **Orchestrator** computes penalty matrix (***P***) (e.g., identity matrix), penalized gradient, and hessian for optimization as

a. 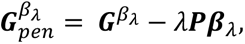
b. 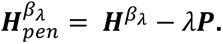
7. **Orchestrator** optimizes and updates iteratively via Newton-Raphson steps (*S*); 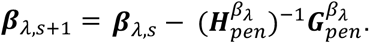
8. **Orchestrator** shares results with participating sites and each site re-runs step-4. Subsequently, steps 5-6 are repeated until convergence |***β****_λ_*_,*s*+1_ − ***β****_λ_*_,*s*_| < *ε* (e.g., *ε* = 1*e* − 4) or maximum number of iterations reached and subsequently enumerate 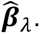
9. **Orchestrator** optimizes the estimation for each value in ***λ*** and obtain 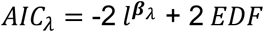 or 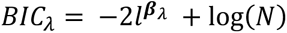 by repeating steps 3-8 for each choice of ***λ***. Define the effective degrees of freedom by 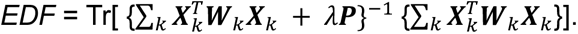
10. **Orchestrator** selects optimum 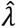 that minimizes *AIC*_***λ***_′*S* or *BIC*_***λ***_′*S*, and compute final estimates 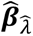 and 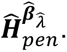

### 4.5 Uncertainty quantification of coefficients

To account for uncertainty in estimation, Fisher information matrix was computed, and the standard errors for ***β*** were obtained. Due to the bias introduced by penalization, Hessian-based standard errors described above may be suboptimal. Consequently, to ensure robust inference, percentile-based bootstrap confidence intervals were calculated based on B runs using a distributed mechanism. Instead of employing a resampling-based bootstrap, a nonparametric approach, such as the fractional random weight bootstrap^22^ (FRWB), was utilized to address the scarcity and sparsity of features across sites, as this method allows for the exploitation of all data rather than resampling with replacement. Alternatively, the Poisson bootstrap (PB) can also be employed. To ensure the reproducibility of weights, the orchestrator shares the seed numbers with sites for all B (e.g., 300 or 1000) runs. Figure SM1 provides technical details of the algorithm to generate bootstrap-based confidence intervals.

## SUPPLEMENTARY MATERIAL

Supplementary material is available along with the submission with additional numerical results pertinent to the study.

## DATA AVAILABILITY

The raw, individual University of Colorado (CU) patient data are protected and not available due to data privacy laws. The processed data are available at reasonable request to the corresponding author. The Flatiron Health data that supported the findings of this study were originated by and are the property of Flatiron Health, Inc., which has restrictions prohibiting the authors from making the data set publicly available. Requests for data sharing by license or by permission for the specific purpose of replicating results in this manuscript can be submitted to.

## STATEMENT OF ETHICS

This retrospective study was approved by University of Colorado internal review board (IRB) and used a limited dataset with a waiver of consent.

