## Supplementary Materials for "Federated penalized piecewise exponential model for horizontally distributed survival data: FedPPEM"

This section provides additional pertinent results related to the study. There are five Supplemental Figures and all of which are referenced in the main body of the paper.

### SUPPLEMENTAL FIGURE LEGENDS

**Supplemental Figure 1. Algorithm of federated penalized piecewise exponential model (FedPPEM) with fractional random weight bootstrap runs.**

**Supplemental Figure 2. Comparison between federated and pooled estimates using FedPPEM and meta-analysis.** Reported are (a) parameter estimates for federated versus pooled, (b) Corresponding standard errors, The results are based on data from 33 sites, each with more than 5 patients, totaling 1,192 patients in the pooled analytical set.

**Supplemental Figure 3. Comparison between federated and pooled estimates using FedPPEM and meta-analysis.** Reported are (a) parameter estimates for federated versus pooled, (b) Corresponding standard errors, The results are based on data from 11 sites, each with more than 20 patients, totaling 980 patients in the pooled analytical set.

**Supplemental Figure 4**. **Bootstrap-specific percentile confidence intervals.** Reported are the 95% lower-bounds (left) and upper-bound (right) based on the FedPPEM and pooled penalized GLM model. Results are based on the sites having more than 50 patients (N = 980 and K = 7).

**Supplemental Figure 5. Approximation between the pooled Cox-PH with L2 norm penalty and pooled penalized GLM with Poisson regression**. Reported are the estimates based on three analytical sets with the number of sites equal to (left) 7, (center) 11, and (right) 33.

**Supplemental Figure 1. Supplemental Figure 1. Algorithm of federated penalized piecewise exponential model (FedPPEM) with fractional random weight bootstrap runs.**

1. Each **site** performs standardization and normalization of features, impute any missing values according to the pre-defined protocol and shares maximum follow-up time. Any deviation to the protocol across sites is identified.
2. **Orchestrator** shares with sites the analytical codes (.R) for implementation, pseudo IDs for site labeling, a grid of potential tuning parameters $\boldsymbol{\lambda}$**,** global knots for time-intervals, total number of patients $N$, and a set of seed numbers (**S**) to enumerate FRWB or PB specific weights ($\tilde{\boldsymbol{f}}_{bk}$) for $b = 1,\ldots,B$ runs
3. Each **site** independently transforms survival response, design matrix, and into piecewise exponential data (PED) format and obtain $\boldsymbol{X}_k$, $\boldsymbol{y}_k$, $\boldsymbol{o}_k$, and $\boldsymbol{f}_{bk}'s$ exploiting dataset with size $n_k$. Let the $b$-th bootstrap weights be $\boldsymbol{F}_{bk} = diag(\boldsymbol{f}_{bk})$.
4. Set initial parameters as $\boldsymbol{\beta}_{b\lambda} = \{0,\ldots,0\}$. For each value in $\boldsymbol{\lambda}$ and for each $b \in B$, every **site** shares the following aggregated statistics with the **orchestrator**-
   a. $(p \times 1)$ vectors of gradients $\boldsymbol{g}_{bk}^{\beta_\lambda} = \boldsymbol{X}_k^T \boldsymbol{F}_{bk}(\boldsymbol{y}_k - \boldsymbol{\mu}_{bk})$ where $\boldsymbol{\mu}_{bk} = \exp(\boldsymbol{\eta}_{bk})$ ; $\boldsymbol{\eta}_{bk} = \boldsymbol{X}_k^T \beta_{b\lambda} + \boldsymbol{o}_k$
   b. $(p \times p)$ matrices of hessians $\boldsymbol{H}_{bk}^{\beta_\lambda} = -\boldsymbol{X}_k^T \boldsymbol{W}_{bk} \boldsymbol{F}_{bk} \boldsymbol{X}_k$ where $\boldsymbol{W}_{bk} = diag(\boldsymbol{\mu}_{bk})$
   c. Scalar values of likelihood $l_{bk}^{\beta_\lambda} = \sum_i \sum_j \{f_{ijbk} y_{ijk} \log(\mu_{ijbk}) - \mu_{ijbk}\}$
5. **Orchestrator** aggregates the site-specific shared data; $\boldsymbol{G}_b^{\beta_\lambda} = \sum_k \boldsymbol{g}_{bk}^{\beta_\lambda}$; $\boldsymbol{H}_b^{\beta_\lambda} = \sum_k \boldsymbol{H}_{bk}^{\beta_\lambda}$; $l_b^{\beta_\lambda} = \sum_k l_{bk}^{\beta_\lambda}$ ; $N = \sum_k n_k$.
6. **Orchestrator** computes penalty matrix ($\boldsymbol{P}$) (e.g., identity matrix), penalized gradient, and hessian for optimization as
   a. $\boldsymbol{G}_{b,pen}^{\beta_\lambda} = \boldsymbol{G}_b^{\beta_\lambda} - \lambda \boldsymbol{P} \boldsymbol{\beta}_{b\lambda}$,
   b. $\boldsymbol{H}_{b,pen}^{\beta_\lambda} = \boldsymbol{H}_b^{\beta_\lambda} - \lambda \boldsymbol{P}$.
7. **Orchestrator** optimizes and updates iteratively via Newton-Raphson steps ($s$); $\boldsymbol{\beta}_{b\lambda,s+1} = \boldsymbol{\beta}_{b\lambda,s} - (\boldsymbol{H}_{b,pen}^{\beta_\lambda})^{-1} \boldsymbol{G}_{b,pen}^{\beta_\lambda}$.
8. **Orchestrator** shares results with sites and each site re-runs step-4. Steps 4-6 are repeated until convergence $|\boldsymbol{\beta}_{b\lambda,s+1} - \boldsymbol{\beta}_{b\lambda,s}| < \epsilon$ (e.g., $\epsilon = 1e-4$) or maximum number of iterations reached and subsequently enumerate $\widehat{\boldsymbol{\beta}}_{b\lambda}$.
9. **Orchestrator** optimizes estimation for each value $\lambda$ for each FRWB or PB runs and obtain $AIC_{b\lambda}$ = -2$l_b^{\beta_\lambda}$ + 2$EDF_b$ or $BIC_{b\lambda} = -2 l_b^{\beta_\lambda} + \log(N)\, EDF_b$ by repeating **steps 3-8** for each choice of $\lambda$. Define the effective degrees of freedom by $EDF_b$ = Tr[ $\boldsymbol{F}_{bk}\{\sum_k \boldsymbol{X}_k^T \boldsymbol{W}_{bk} \boldsymbol{X}_k + \lambda \boldsymbol{P}\}^{-1} \{\sum_k \boldsymbol{X}_k^T \boldsymbol{W}_{bk} \boldsymbol{X}_k\}$].
10. **Orchestrator** selects $\hat{\lambda}_b$ that minimizes $\boldsymbol{AIC_{b\lambda}}$ or $\boldsymbol{BIC_{b\lambda}}$ and computes final estimates $\widehat{\boldsymbol{\beta}}_{b\hat{\lambda}_b}$ for the $b$-th run.
11. Obtain percentile-based (e.g., 2.5th and 97.5th) bootstrap confidence intervals for $\widehat{\boldsymbol{\beta}}_{\hat{\lambda}}$.

**Supplemental Figure 2. Comparison between federated and pooled estimates using FedPPEM and meta-analysis.** Reported are (a) parameter estimates for federated versus pooled, (b) Corresponding standard errors, The results are based on data from 33 sites, each with more than 5 patients, totaling 1,192 patients in the pooled analytical set.

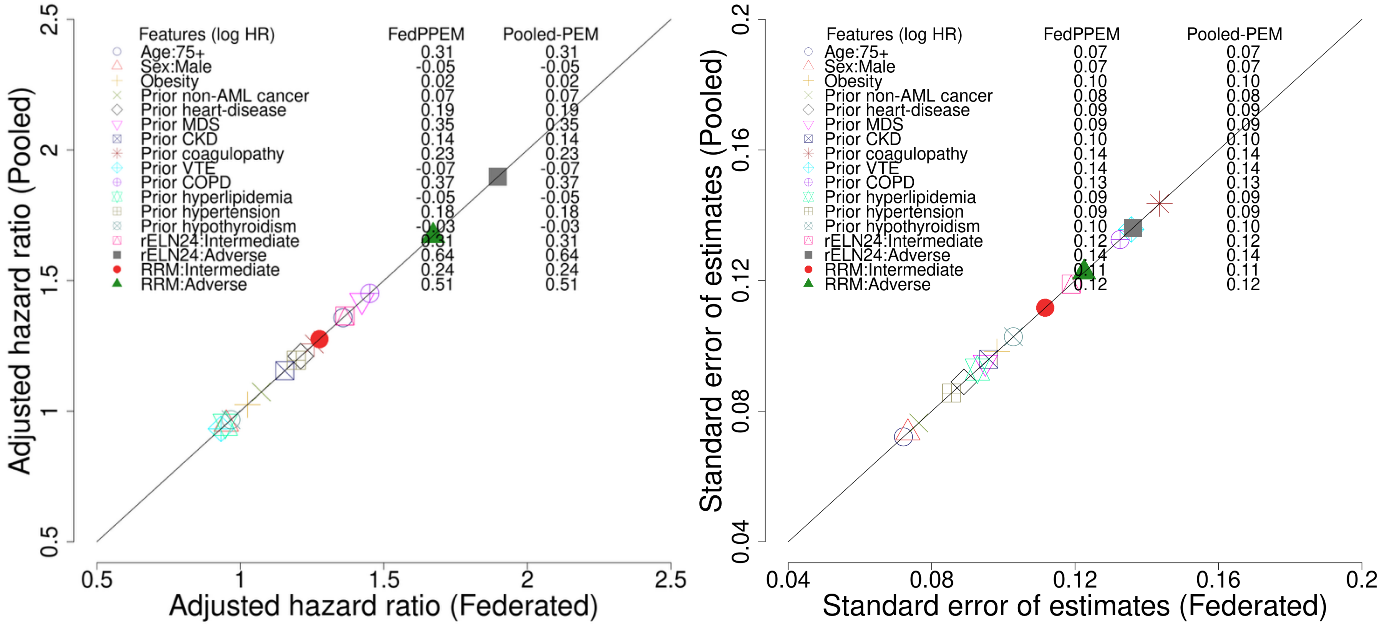

**Supplemental Figure 3. Comparison between federated and pooled estimates using FedPPEM and meta-analysis.** Reported are (a) parameter estimates for federated versus pooled, (b) Corresponding standard errors, The results are based on data from 11 sites, each with more than 20 patients, totaling 980 patients in the pooled analytical set.

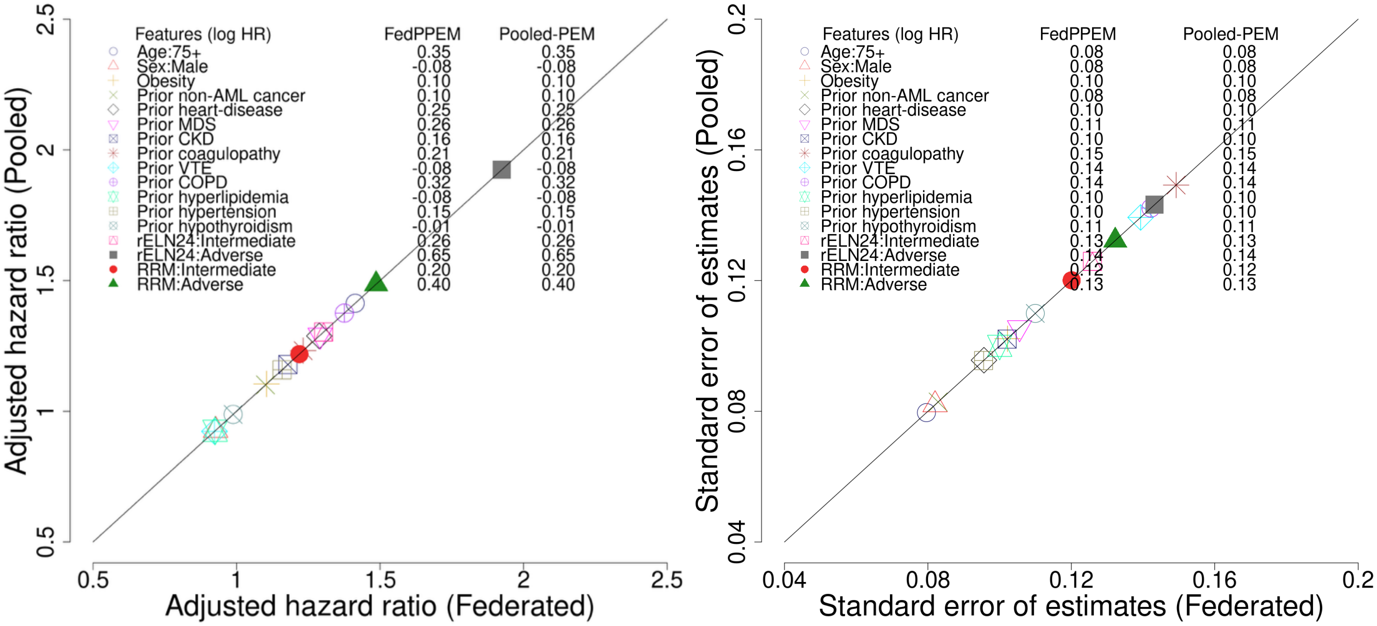

**Supplemental Figure 4**. **Bootstrap-specific percentile confidence intervals.** Reported are the 95% lower-bounds (left) and upper-bound (right) based on the FedPPEM and pooled penalized GLM model. Results are based on the sites having more than 50 patients (N = 980 and K = 7).

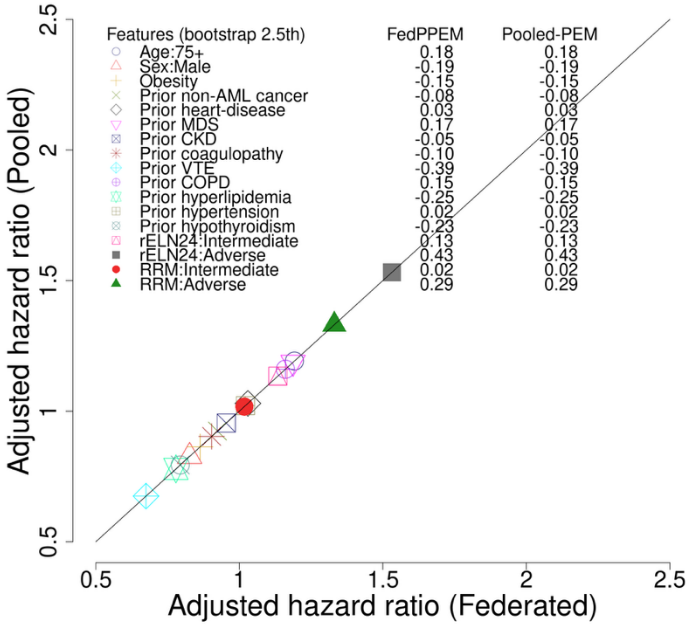


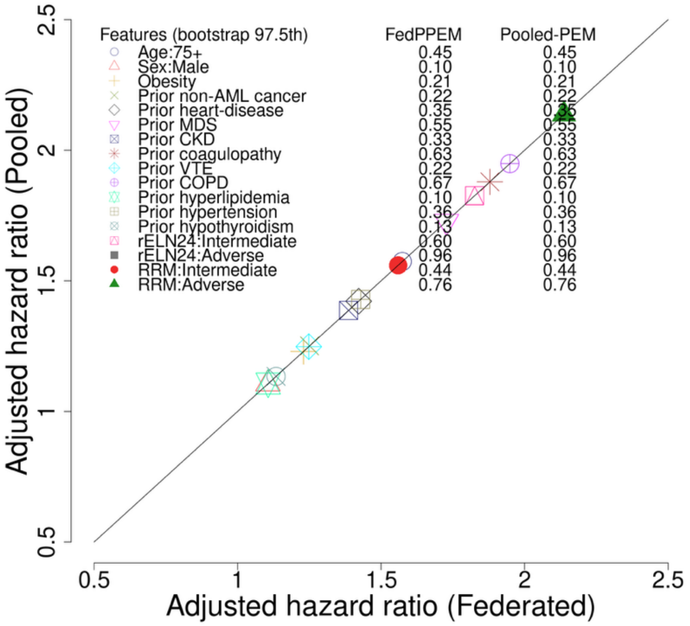

**Supplemental Figure 5. Approximation between the pooled Cox-PH with L2 norm penalty and pooled penalized GLM with Poisson regression**. Reported are the estimates based on three analytical sets with the number of sites equal to (left) 7, (center) 11, and (right) 33.

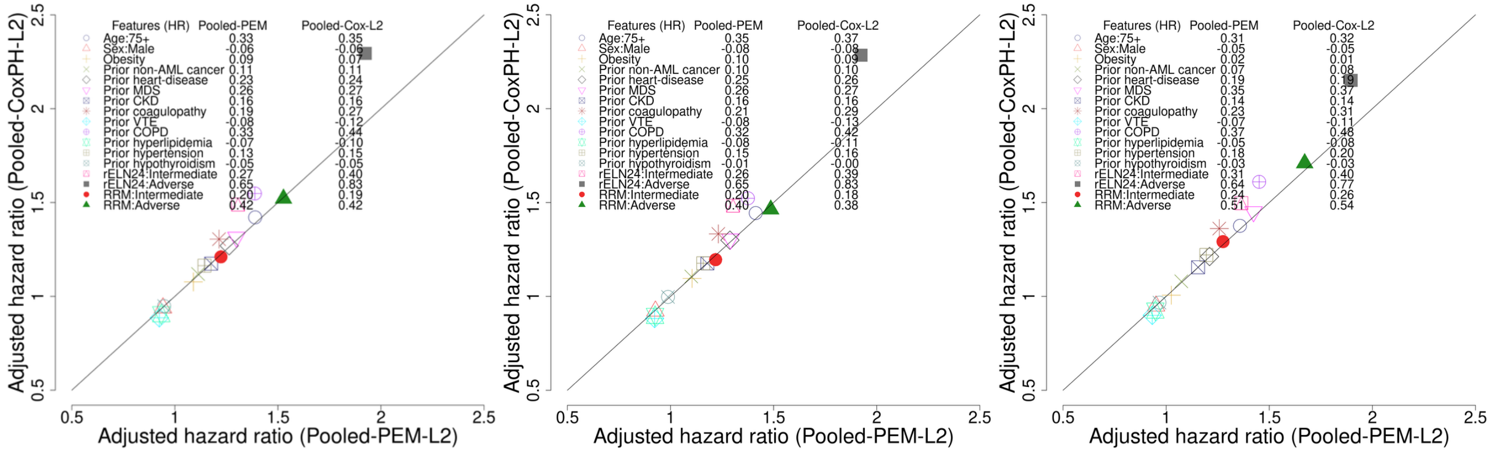
